# Evaluation of machine learning for predicting COVID-19 outcomes from a national electronic medical records database

**DOI:** 10.1101/2022.04.13.22273835

**Authors:** Sean Browning, Scott H. Lee, Ermias Belay, Jennifer DeCuir, Shana Godfred Cato, Pragna Patel, Noah Schwartz, Karen K. Wong

## Abstract

**Objective:** When novel diseases such as COVID-19 emerge, predictors of clinical outcomes might be unknown. Using data from electronic medical records (EMR) allows evaluation of potential predictors without selecting specific features *a priori* for a model. We evaluated different machine learning models for predicting outcomes among COVID-19 inpatients using raw EMR data.

**Materials and Methods:** In Premier Healthcare Data Special Release: COVID-19 Edition (PHD-SR COVID-19, release date March, 24 2021), we included patients admitted with COVID-19 during February 2020 through April 2021 and built time-ordered medical histories. Setting the prediction horizon at 24 hours into the first COVID-19 inpatient visit, we aimed to predict intensive care unit (ICU) admission, hyperinflammatory syndrome (HS), and death. We evaluated the following models: L2-penalized logistic regression, random forest, gradient boosting classifier, deep averaging network, and recurrent neural network with a long short-term memory cell.

**Results:** There were 57,355 COVID-19 patients identified in PHD-SR COVID-19. ICU admission was the easiest outcome to predict (best AUC=79%), and HS was the hardest to predict (best AUC=70%). Models performed similarly within each outcome.

**Discussion:** Although the models learned to attend to meaningful clinical information, they performed similarly, suggesting performance limitations are inherent to the data.

**Conclusion:** Predictive models using raw EMR data are promising because they can use many observations and encompass a large feature space; however, traditional and deep learning models may perform similarly when few features are available at the individual patient level.

## Background and objective

Predicting clinical outcomes traditionally involves abstracting clinical features from the medical record that may be relevant to the outcomes of interest. However, this approach requires relevant features to be specified *a priori*. For emerging diseases, the relevant features may be unclear. Rich clinical data are often reduced to simpler elements; for example, vasopressor use may be simplified to “yes/no” rather than capturing the type of vasopressor, the dose over time, and how its use relates to the time course of other medications or laboratory values. Ideally, all of a patient’s health data could be included in a predictive model, without discarding information that was not in a predetermined set of features.

Previously, predicting clinical outcomes using electronic medical records (EMR) data was often unfeasible due to data inaccessibility, prohibitive data licensing costs, resource constraints for data processing and storage, and the technical complexity of analysis. More EMR data are becoming available, however, and methods to analyze them have improved, allowing for high-performing predictive models using raw, often unstructured, data. The infrastructure costs of analysis at scale, like cloud computing and storage, have plummeted, and research groups at the forefront of EMR analysis often make their code, data, and pretrained models openly available. The result is that scientists can build predictive models for a wide variety of applications, including prognosis and diagnosis [1] medication recommendation [2], inpatient management [3], and even synthetic data generation [4,5].

Beyond these applications, rich patient data in EMRs also allows study of rare outcomes without resource-intensive case-control studies. Routinely collected data offer promise for hypothesis generation and early exploration of associations where a mechanistic understanding is of lesser importance, and in many cases they can support development of clinically-meaningful prediction tools [6].

Many advances in EMR analysis have been driven by developments in deep learning, which allows modeling of complex dependencies between predictors and outcomes, even when those dependencies are spread across time (e.g., a history of cardiovascular disease informing severe outcomes in COVID-19 hospitalizations) or across heterogenous, unstructured datatypes (e.g., free-text physician notes informing a clinical diagnosis of Parkinson’s disease). Although these models are often time-intensive to build, especially when there is no existing data preprocessing pipeline tailored to the structure of a particular vendor’s EMR, they take far less time to train, validate, test, and deploy than longitudinal studies do to design, approve, and conduct. Because they are relatively straightforward to update as new training data become available, they are also much easier to change in response to emerging conditions.

We evaluated five machine learning models for their potential to predict clinical outcomes among COVID-19 patients from raw EMR data. For our outcome of interest, we defined a hyperinflammatory syndrome (HS) as a proxy for multisystem inflammatory syndrome in adults (MIS-A), a rare complication of COVID-19 characterized by a severe inflammatory phenotype for which relatively little is known about risk factors and predictors [7]. We also examined model performance for predicting intensive care unit (ICU) admission and death. A secondary goal was to explore the challenges of working with EMR data at scale, which we hope will be a useful point of reference for practitioners undertaking similar projects.

## Prior Work

### Prediction tasks with EMRs

Machine learning models have been widely explored for solving prediction tasks with EMR data. Many studies have explored general uses, including representation learning for medical concepts [4], automated ICD coding [8], medication recommendation [2], diagnosis [1,9], and prognosis [10,11]. Other studies have focused more narrowly on predicting outcomes for specific conditions, such as acute kidney injury (AKI) [3] and heart failure [12]. Some [3] were developed with clinical deployment in mind, but many were developed to explore ways of handling the size and richness of raw EMRs [1].

With respect to COVID-19, much recent work on machine learning and EMRs has focused on predicting specific outcomes, including ICU transfer [13], intubation [14], and death [6,15,16]. Some models make predictions for a single time point, like the end of a stay in the ICU, while others make predictions across a sliding window of time points, like for each day of the same stay. The sliding window approach requires time-aware, often deep models, which tend to be more data-hungry and computationally complex than single-output models. Because training data are limited for rare outcomes such as MIS-A, we focus on the task of predicting visit-level outcomes.

### Data representations for EMRs

Because raw EMR data tend to be rich, with a high-dimensional feature space, and long, with observations being ordered over long periods of time, finding a data representation that is computationally efficient but informative with respect to the outcomes is challenging. To address the problem of high-dimensionality, studies generally take one of two approaches: manual feature selection and engineering before model training [13]; and automatic feature representation learning during model training [1]. Because manual feature engineering takes time and expertise, and automatic representation learning has the potential to discover more efficient feature representations than what could be crafted by hand, we chose the latter. Similarly, to address the problem of longitudinality, we experimented with two approaches: flattening observation sequences before modeling, and modeling observation sequences directly, e.g., with recurrent neural networks (RNNs).

### Model types for EMRs

Model types used with EMRs tend to follow the chosen data representation. With raw features, models tend to be based on deep neural networks, which are generally more adept at learning high-level feature representations than traditional models like support vector machines (SVM), random forests (RF), or regularized generalized linear models (GLM). The same holds for time-aware or otherwise sequential features, which are typically handled by attention-based or RNNs that can extract information from the order, rather than just the presence or absence, of specific features. Popular architectures in recent works include versions of the Transformer [17] and the long short-term memory (LSTM) RNN [18], both of which are capable of feature-level representation learning and outcome-level prediction from time-ordered observations in raw EMRs.

When the feature space is small, or when the time dimension is removed, traditional models often work well. Many of these models, especially tree-based methods like RF or gradient-boosting machines (GBMs), are also more directly interpretable than deep neural networks (recent advances in attribution methods notwithstanding [19]), and so they are often used as an initial step. For emerging diseases and syndromes like COVID-19 and MIS-A, respectively, strong interpretability is often more easily transferrable to symptom-based case definitions that can be used in low-resource clinical settings for screening, diagnosis, and surveillance. For these reasons, we explored three combinations of data representations and model types: a non-sequential data representation with shallow models (SVM, RF, GBM, and GLM); a non-sequential data representation with a deep model (a deep averaging network, or DAN) [20]; and a time-ordered data representation with a deep sequential model (LSTM).

### Heuristics for building end-to-end prediction pipelines

Taken together, a time-ordered data representation and deep sequential model represent a theoretical approach for learning from raw EMRs: they preserve the full richness of each patient’s clinical time course, and they are capable of learning meaningful higher-level representations from feature spaces that are large and often sparse. Deep sequential models can be resource-intensive to train and deploy, especially when using large, attention-based networks like the Transformer, and the technical effort required to develop bespoke data preprocessing pipelines from raw EMRs can be immense. It is reasonable to ask is when to use this approach instead of selecting or engineering features by hand and then using less resource-intensive models to make the predictions. Considerations include the availability of technical resources, like engineers for constructing the data pipelines and Graphics Processing Units (GPUs) for training deep neural networks, and the characteristics of the raw data themselves. Our study touches on several of these factors, and our results show that the theoretically ideal approach may not always be the most reasonable or effective.

## Materials and Methods

### Data Sources

The PHD-SR COVID-19 dataset includes billing codes, laboratory tests ordered, laboratory results, vital signs, and ICD-10 diagnosis and procedure codes. Data were linkable by visit and patient identifiers.

While patient identifiers usually represent unique individuals, they are not harmonized across health systems participating in the Premier network, and so it is possible that some patients are represented by more than one identifier, distributing their true medical histories across multiple visit-sequences.

### Inclusion Criteria

We included patients with at least one inpatient visit with a primary or secondary COVID-19 ICD-10 code with admission dates during February 1, 2020 through April 30, 2021 and defined the first COVID-19 inpatient visit as the target visit. For admissions beginning February 1 to April 1, 2020 and ending March 1 through April 1, 2020, we used the working ICD-10 code B97.2, “Coronavirus as the cause of diseases classified to other chapters.” For admissions after April 1, 2020, we used the emergency use COVID-19 diagnosis code U07.1, ”COVID-19, virus identified.” With potential target visits identified, we then excluded patients aged <18 years at their target visit. For the remaining patients, we used all available inpatient and outpatient visits after January 1, 2018 but prior to their target visit to construct their clinical timelines.

### Outcome Definitions

Most of the data comprised billing and procedure codes and selected laboratory results, making implementation of the MIS-A case definition, which relies on several clinical criteria, unfeasible [21]. As a proxy for MIS-A, we defined a hyperinflammatory syndrome (HS), which likely includes MIS-A as well as severe manifestations of acute COVID-19 (Appendix). ICU admission was defined by presence of billing codes for Room & Board ICU or Room & Board Stepdown during a hospitalization for COVID-19. The death outcome was defined as death from any cause occurring during the first COVID-19 hospitalization.

### Feature Tokenization and Data Model

Our data model is similar to the model used by Rajkomar et al. (2018). First, we discretized all continuous features associated with laboratory results and vital signs by computing sample quantiles and appending a text label. Then we transformed all discrete feature values into short tokens, such as descriptive billing and laboratory order codes, and we assigned any visit-level codes that did not have timing information to the last day of the visit. We then used a text vectorizer to convert the groups of tokens into lists of integers representing the unigrams in the sequence. To control the overall vocabulary size, we set the vectorizer’s minimum document frequency to 5 (i.e., we omitted any code appearing fewer than 5 times across all patients), yielding a total vocabulary of 34,422 unique codes.

For each patient, we aggregated all codes and discretized values into “bags” representing a single day of their available medical history, and we arranged the resulting list of lists by day relative to the index date for their corresponding medical record ID. Because some patients had many (in some cases, several hundred) visit-days recorded, we used a lookback period of 225 days (which comprised most patients’ full histories) to trim the sequences for modeling, ending each sequence on the first day of the first inpatient COVID-19 visit. This cutoff (24 hours) became the pre-diction horizon.

For the LSTM, the input for a single patient was this complete sequence of bags of features, and for all other models, it was their unordered concatenation. To determine how well we could predict outcomes from the target visit alone, we also discarded medical history and considered models trained on features gathered from only the first day of the inpatient COVID-19 visit.

We separately encoded patient demographic features at the patient level as tokens and combined them with the list of day-level features. These included: gender, ethnicity, race, and age (discretized into sample deciles). For the LSTM model, we embedded these features, passed them through a fully connected layer, and concatenated the resulting vector to the sequence of vectors produced by the recurrent cell before classification by the final layer. For all other models, as before, we added the demographic features for each patient to their existing list of codes.

### Models

We produced two deep learning models using Keras [22] with the TensorFlow backend: a RNN with an LSTM cell, and a DAN.

The LSTM model contained three embedding layers: the first two embed the visit-level features into an output dimension of 64 and 1, and the third embeds the patient-level demographics. We multiplied the output tensors of the visit-level embedding layers and computed the mean along the second axis. We fed this average into an LSTM recurrent layer with 128 neurons and concatenated the output with the output of the patient-level embeddings. The final dense layer contained a single neuron with a sigmoid activation for prediction.

Both models trained for 20 epochs with a batch size of 64, with early stopping on a validation loss criterion if the model failed to improve in 3 epochs. We used the Adam [23] algorithm for optimization with a learning rate of 0.001, and we used binary cross-entropy for the loss function.

In addition to the deep models, we also trained classical ML models, including an L2-regularized logistic regression, a RF [24], and a gradient boosting classifier [25]. We implemented these models in scikit-learn [26] and trained them with their default hyperparameters unless otherwise noted.

### Evaluation Metrics

We split patients into training (64%), test (20%), and validation (16%) sets using multilabel stratified random sampling on the outcome. We evaluated all models based on a variety of common classification metrics, including sensitivity, specificity, F1-score, area under the receiver operating characteristic curve (AUC), and Brier score. For inference, we computed 95% bias-corrected and accelerated bootstrap confidence intervals [27]. We did not adjust the intervals for multiplicity, and we did not perform direct pairwise comparisons between models.

### Software

We used R 4.1.0 [28] and SQL for data extraction and preprocessing, and we used Python 3.8 for feature extraction, modeling, and statistical analysis. Baseline models were built in scikit-learn, and deep models were built using Keras [22] with the TensorFlow 2.0 backend [29]. Our code is available at github.com/cdcai/premier_analysis.

### Ethics Statement

The Institutional Review Board of the Centers for Disease Control and Prevention waived ethical approval for this work because the disclosed PHD-SR COVID-19 data are considered de-identified.

## Results

### Data Characteristics

Table 1 shows the distribution of the feature vocabulary *V* across clinical data categories with the patient-level distributions of unique features for different time periods (i.e., all visits vs. the first 24 hours of the target inpatient visit). The feature space is large (34,422), especially relative to the median number of unique features recorded per patient (96). Most patients have relatively little information available about vitals and microbiological lab results. Among patients with more than one visit-day in the dataset, most information is contained by the first 24 hours of the target inpatient visit.

**Table 1.**
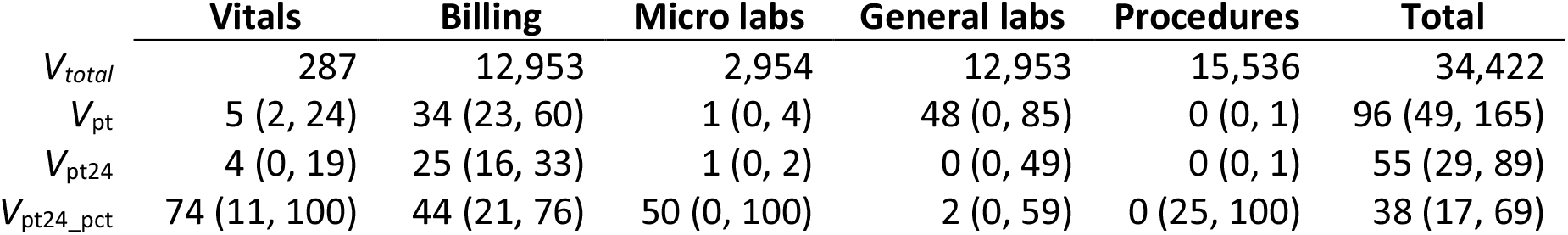
Number of unique features for each of our 5 source data tables in total (V_total_), followed by the median and interquartile ranges for the numbers of unique features per table by patient from all visits (V_pt_) and from only the first 24 hours of the target inpatient visit (V_pt24_). The final row (V_pt24_pct_) shows the median and interquartile ranges for the percent of patient-level unique codes present during the first 24 hours of the target visit but absent from all of the previous visit-days.

Figure 1 shows the distribution of patient-level visit sequence lengths. Although some patients had many days of follow-up available for modeling, most did not, with the median number of days being 1 or 2 for most outcomes. Most patients only had a few recorded interactions with Premier network facilities, so the amount of information available for any single patient was small.

**Figure 1.**
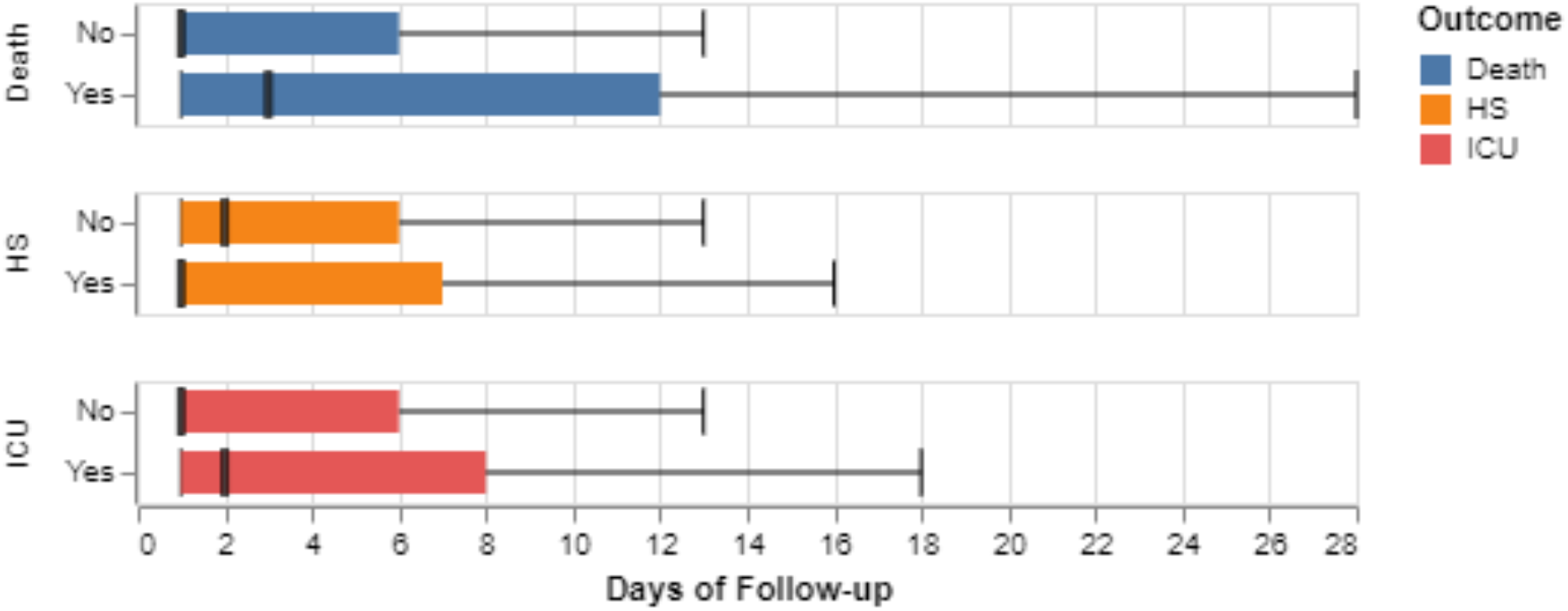
Box plot showing the interquartile range (IQR) and median for patient-level sequence lengths by outcome.

Our dataset contained 57,355 patients in total. After the stratified train-test-validate split, our test dataset included 11,471 patients, of which 3,211 (28%) met our criteria for ICU admission, 1,241 (11%) for HS, and 1,300 (11%) for death.

### Classification Performance

Metrics for all models and tasks are shown in Table 2. Model AUCs were 70% for predicting HS, 77% for predicting death, and 79% for predicting ICU admission. F1-scores were substantially lower for the death (33%) and HS (29%) tasks than for the ICU task (55%). Although this may be partially due to differences in class imbalance (about three times as many patients were admitted to the ICU as died or developed HS), it likely reflects a true increase in difficulty in predicting those outcomes. Although Brier scores for all models were low, histograms for these two outcomes also showed a large degree of overlap between the distributions of scores for negative patients and positive patients (Supp. Fig 1), suggesting a relatively high Bayes error rate for these tasks. Overall, models were competitive with each other across the metrics we analyzed. The similarity in performance across models can also be seen in the ROC curves shown in Figure 2, where the model-specific true positive rates (TPRs) are similar at most points in the space.

**Table 2.** Point estimates and 95% confidence intervals for sensitivity (sens), specificity (spec), positive predictive value (ppv), negative predictive value (npv), F1-score (f1), Brier score (brier), and area under the receiver operating characteristic curve (auc) for all models across our 3 outcomes: death, hyperinflammatory syndrome (HS), and ICU admission. Top values for each metric/task combination are shown in bold.

|  |  | sens | spec | ppv | npv | f1 | brier | auc |
| --- | --- | --- | --- | --- | --- | --- | --- | --- |
| **Death** |  |  |  |  |  |  |  |  |
|  | lgr | 32 (30, 35) | **91 (90, 91)** | **27 (25, 29)** | 93 (93, 93) | 29 (28, 32) | **8 (8, 8)** | 75 (74, 77) |
|  | rf | 43 (41, 46) | 86 (86, 87) | 24 (22, 26) | **94 (93, 94)** | 31 (30, 33) | 8 (7, 8) | 75 (74, 77) |
|  | gbc | 41 (38, 43) | 88 (87, 88) | 25 (23, 27) | **94 (93, 94)** | 31 (29, 33) | 8 (7, 8) | **77 (75, 78)** |
|  | dan | 49 (46, 52) | 84 (83, 85) | 24 (22, 26) | **94 (94, 95)** | 32 (30, 34) | 8 (7, 8) | **77 (76, 79)** |
|  | lstm | **49 (46, 51)** | 85 (84, 85) | 25 (23, 26) | **94 (94, 95)** | **33 (31, 34)** | 8 (7, 8) | 76 (75, 77) |
| **HS** |  |  |  |  |  |  |  |  |
|  | lgr | 41 (38, 43) | 80 (80, 81) | 19 (18, 21) | 92 (92, 93) | 26 (24, 28) | **9 (9, 10)** | 67 (65, 68) |
|  | rf | 46 (44, 49) | 80 (79, 81) | **21 (19, 22)** | **93 (92, 93)** | **29 (27, 30)** | **9 (8, 9)** | 69 (68, 70) |
|  | gbc | **48 (45, 50)** | 79 (79, 80) | **21 (19, 22)** | **93 (93, 93)** | **29 (27, 30)** | **9 (8, 9)** | **70 (69, 71)** |
|  | dan | 42 (40, 45) | **81 (81, 82)** | 20 (19, 22) | **93 (92, 93)** | 27 (26, 29) | **9 (8, 9)** | 69 (68, 71) |
|  | lstm | 45 (43, 48) | 80 (79, 81) | 20 (19, 21) | **93 (92, 93)** | 28 (27, 29) | **9 (8, 9)** | 69 (68, 70) |
| **ICU** |  |  |  |  |  |  |  |  |
|  | lgr | **72 (70, 73)** | 71 (70, 72) | 44 (43, 45) | 89 (88, 90) | **55 (54, 56)** | 15 (15, 16) | 78 (77, 79) |
|  | rf | 66 (65, 68) | 74 (73, 75) | 45 (43, 46) | 87 (87, 88) | 54 (53, 55) | 15 (15, 16) | 77 (76, 78) |
|  | gbc | 70 (69, 72) | 71 (70, 72) | 44 (42, 45) | 88 (87, 89) | 54 (53, 55) | 15 (15, 16) | 77 (77, 78) |
|  | dan | 71 (69, 72) | 73 (72, 73) | **45 (44, 46)** | **89 (88, 89)** | **55 (54, 56)** | **15 (15, 15)** | **79 (78, 79)** |
|  | lstm | 62 (60, 63) | **76 (75, 77)** | **45 (44, 46)** | 86 (85, 87) | 52 (51, 53) | 16 (15, 16) | 75 (74, 76) |

**Figure 2.**
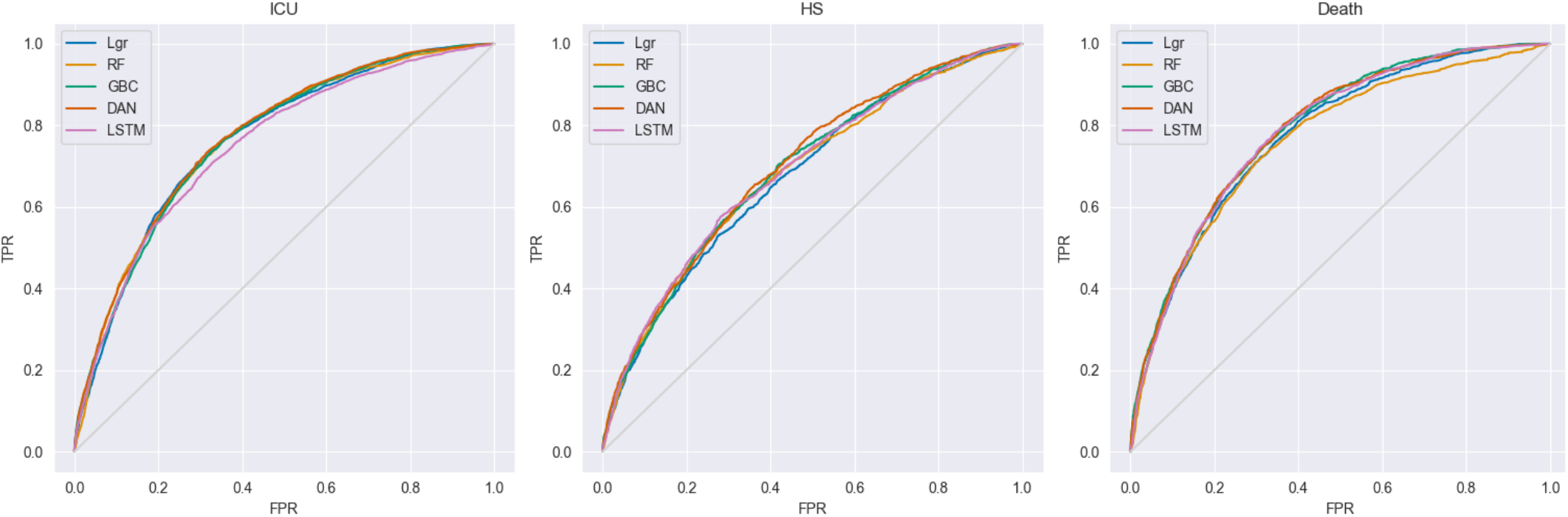
Receiver operating characteristic (ROC) curves for our models in predicting the three outcomes: ICU admission, hyperinflammatory syndrome (HS), and death. The models included a L2-penalized logistic regression (Lgr), a random forest (RF), a gradient boosting classifier (GBC), a deep averaging network (DAN), and a recurrent neural network (RNN) with a long short-term memory (LSTM) cell.

### Model Interpretability

We used three of our baseline models to evaluate whether specific predictors were driving classification performance. From the logistic regression model, we examined the exponentiated coefficients (Supp. Figure 2, github.com/cdcai/premier_analysis/blob/master/lgr_coefs.xlsx), and from both the RF and gradient boosting classifiers, we examined feature importances (github.com/cdcai/premier_analysis/blob/master/importances.xlsx). For the death and ICU outcomes, the models identify several features characteristic of severe acute COVID-19, such as use of resuscitation fluids, vasopressors, and intubation-related medications. The model also identified features related to severe acute disease for the HS outcome, and several were related to laboratory tests (for example, lactate, troponin, and anion gap).

## Discussion

To predict an imprecise clinical outcome like MIS-A, it should be helpful to use raw, rich data and deep models because these are likely to better capture nuanced interactions that are important for the prediction task. High-quality structured clinical data in the early stages of an outbreak may be limited, but EMRs can offer large amounts of data that may be rich, which makes them an attractive data source to evaluate an emerging disease. To predict outcomes among COVID-19 inpatients from our data, we replicated a technique to generate sequences from raw EMRs that can capture aspects of patient history and timeline, in addition to modeling complex within-visit dependencies between several types of clinical information, such as vital signs, laboratory results, and medications received.

This study shows that not all EMR data are equally suitable for this kind of modeling approach. Although we were aware of gaps in data completeness before embarking on our analysis, we were uncertain how detrimental those gaps would be to model performance. Issues with data completeness may not be immediately apparent to analysts approaching a large data set, especially when a preprocessing pipeline for the specific EMR vendor is unavailable. Although our dataset’s feature space was expansive, comprising billing codes, diagnosis codes, laboratory results, and vital sign measurements, it became substantially shallower at the patient level, with each visit containing only a few of the features potentially available.

Interestingly, although the feature space was larger than one might expect from a traditional cohort analysis, it was an order of magnitude smaller than for comparable studies using machine learning with EMR data (for example, Tomasev et al. 2019 cite about 620,000 features, and Rajkomar et al. 2018 cite around 140,000). Whereas vital signs and microbiological results were available for some patients in our cohort, most were available only for the target inpatient visits. Additionally, our data lacked free text in the form of clinical narrative reports, which may have provided a deeper insight into clinical course and trajectory than billing and procedure codes alone. Given recent advances in large-scale language models, like those built from the Transformer [17], we believe this would improve model performance and recommend model developers obtain free-text clinical notes when feasible.

We originally hypothesized that our time-aware model would outperform our baselines and time-agnostic DAN model. Indeed, time-aware models have previously achieved some of the best performance on EMR prediction tasks [1], likely because of the rich health information preserved by intact patient-level time courses. Our results indicate that the time-aware RNN model was not superior to either the baseline or DAN models. Most of the cohort had no additional visit data preceding the prediction horizon. We suspect this was due to the particular blend of provider types (e.g., outpatient vs. inpatient) represented in the data available to us for the analysis. Regardless of the the cause, the lack of time-distributed data limited the performance of our time-aware LSTM model, bringing it on par with both the DAN and the baseline models. These results emphasize that traditional machine learning models may indeed perform as well as deep learning models with certain data.

Instead of modeling the raw EMR data directly, another approach would have been to focus on the selection and engineering of a few key features, either with automated feature selection techniques as in Gao et al. 2020, or by hand. Such an approach could outperform the one demonstrated here, particularly when paired with subject matter expertise. However, *a priori* knowledge of relevant features for an emerging clinical syndrome like MIS-A may not be readily available, and naïve feature engineering and selection over an ample feature space may prove intractable given resource constraints. Ultimately, practitioners must weigh the cost of incorporating domain knowledge where it is available with the potential value added by that approach.

## Conclusion

In this study, we provide a systematic evaluation of different modeling approaches for use on raw EMR data, and we demonstrated the technical complexity of the upstream data pre-processing required to build models on top of those data. The lessons learned from this evaluation can inform future efforts on using EMR and administrative data for an emerging syndrome when more structured data may be limited or when prospective studies are not feasible.

## Data Availability

All data produced are available from Premier Inc. as the PINC-AI Healthcare Data Special Release: COVID-19.

https://offers.premierinc.com/PINC-AI-Healthcare-Data-COVID-Whitepaper-LandingPage.html

## Acknowledgements

We thank Sapna Bamrah Morris for helpful input in study conception.

## Disclaimer

The findings and conclusions in this report are those of the authors and do not necessarily represent the official position of the Centers for Disease Control and Prevention (CDC).

### Appendix: Hyperinflammatory illness case definition

Cases of hyperinflammatory illness were defined by meeting criteria 1, 2, and 3.

1. Hospitalization in a person ≥18 years old, where
  a. for admissions in February-April 2020 and discharges in March-April 2020, there is a primary or secondary ICD-10 code of B97.29: Other coronavirus as the cause of diseases classified elsewhere
  b. for discharges after April 2020, there is a primary or secondary ICD-10 code of U07.1: COVID-19
2. At least 1 of the following markers of inflammation:
  a. IL-6 ≥ 46.5 pg/mL
  b. C-reactive protein ≥ 30 mg/L
  c. Ferritin ≥ 1500 ng/mL
  d. Erythrocyte sedimentation rate ≥ 45 mm/hour
  e. Procalcitonin ≥ 0.3 ng/mL
3. At least 2 of the following categories of clinical complications
  a. Cardiac, defined by at least one of the following ICD-10 codes
    i. I30: Acute pericarditis
    ii. I40: Infective myocarditis
    iii. I25.41: Coronary artery aneurysm
  b. Shock or hypotension, defined by at least one of the following
    i. Systolic blood pressure <90 mmHg on at least 2 days
    ii. At least one of the following ICD-10 codes:
      1. R57, Shock, not elsewhere classified (includes all subcodes)
      2. R65.21, Severe sepsis with septic shock
      3. I95, Hypotension shock (includes all subcodes)
    iii. Vasopressor use on at least 2 days
  c. Gastrointestinal, defined by at least one of the following ICD-10 codes
    i. R10.0, Acute abdomen
    ii. R19.7, Diarrhea, unspecified
    iii. A09, Infectious gastroenteritis and colitis, unspecified
    iv. A08.39, Other viral enteritis
    v. A08.4, Viral intestinal infection, unspecified
    vi. R11.2, Nausea with vomiting, unspecified
    vii. R11.10, Vomiting, unspecified
  d. Thrombocytopenia or elevated D-dimer
    i. Thrombocytopenia, defined by one of the following ICD-10 codes
      1. D69.6, Thrombocytopenia, unspecified
      2. D69.59, Other secondary thrombocytopenia
    ii. D-dimer ≥ 1200 FEU

## Supplemental Figures

**Supplemental Figure 1.**
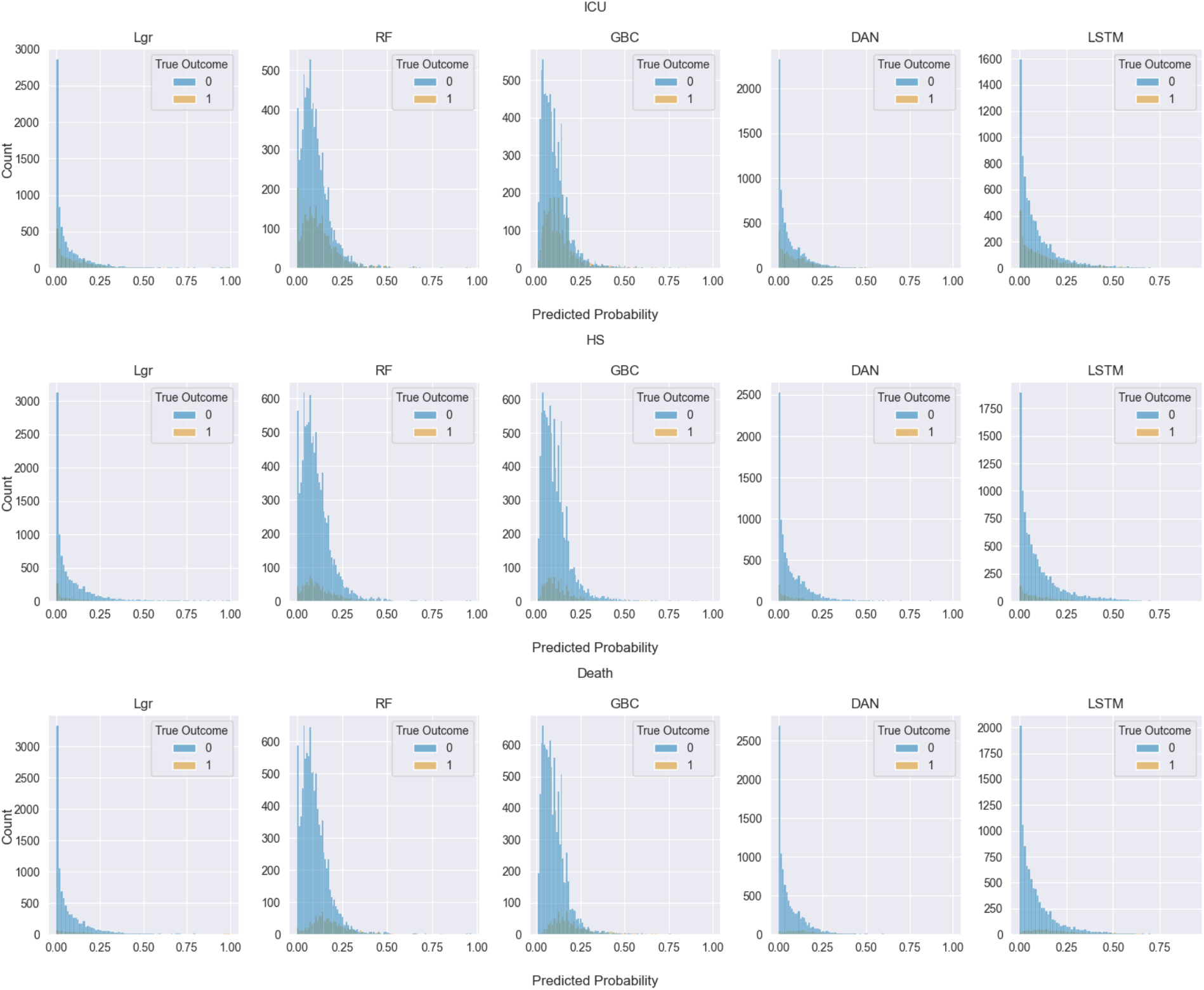
Histograms for the model-derived predicted probabilities for each outcome. Distributions are colored according to the true label. Models are an L1-regularized logistic regression (LGR), random forest (RF), gradient boosting classifier (GBC), deep averaging network (DAN), and RNN with a long-short term memory cell (LSTM).

**Supplemental Figure 2.**
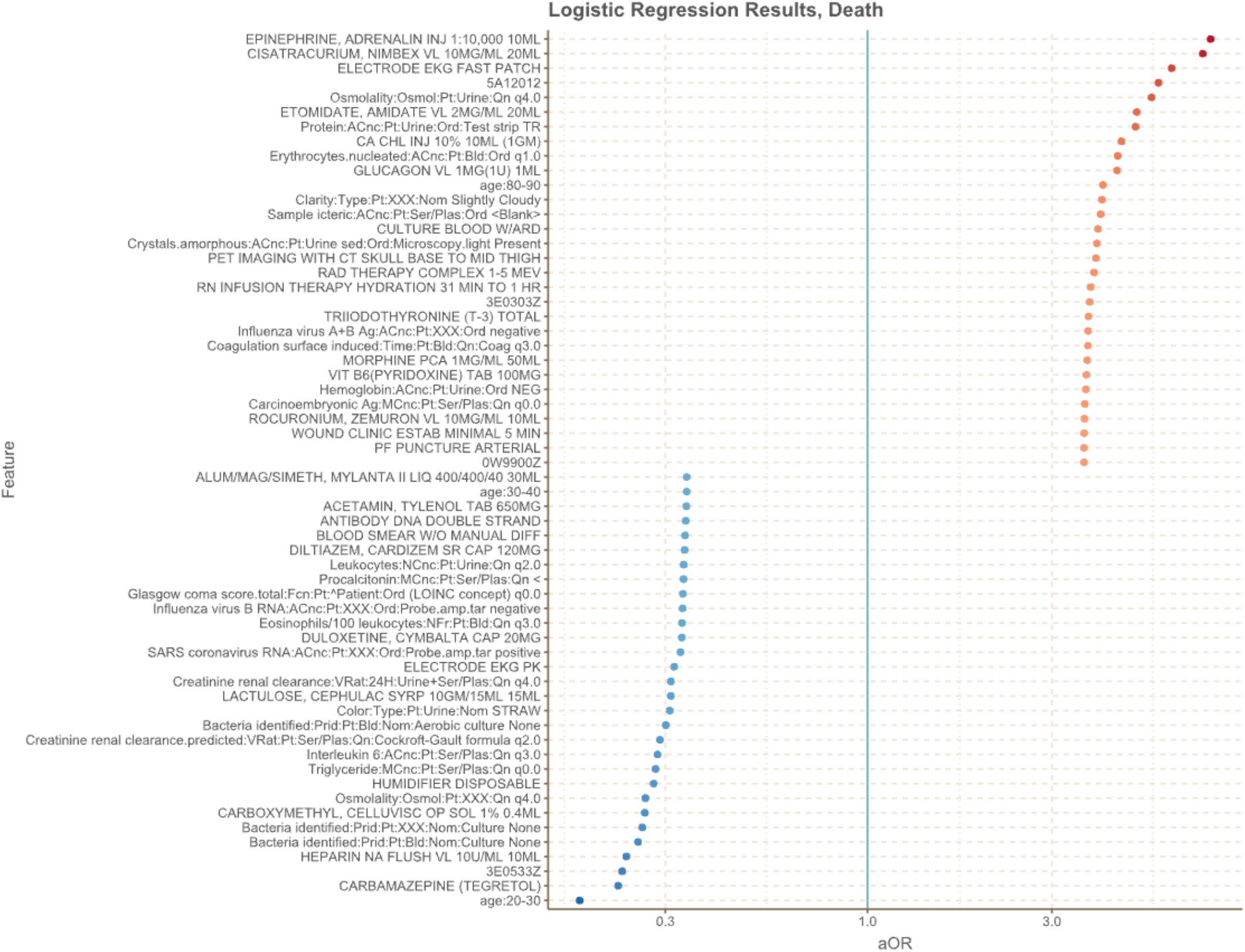
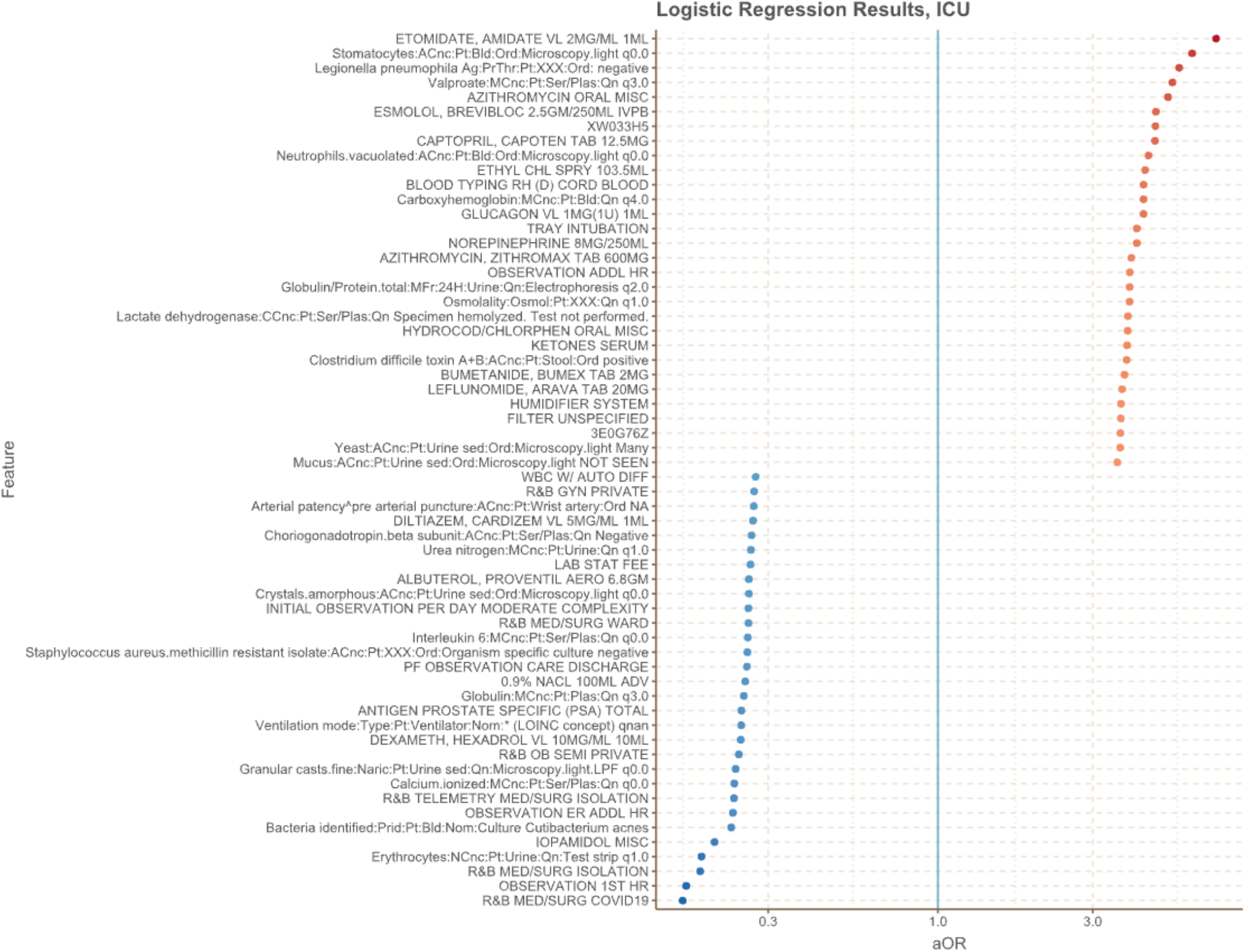
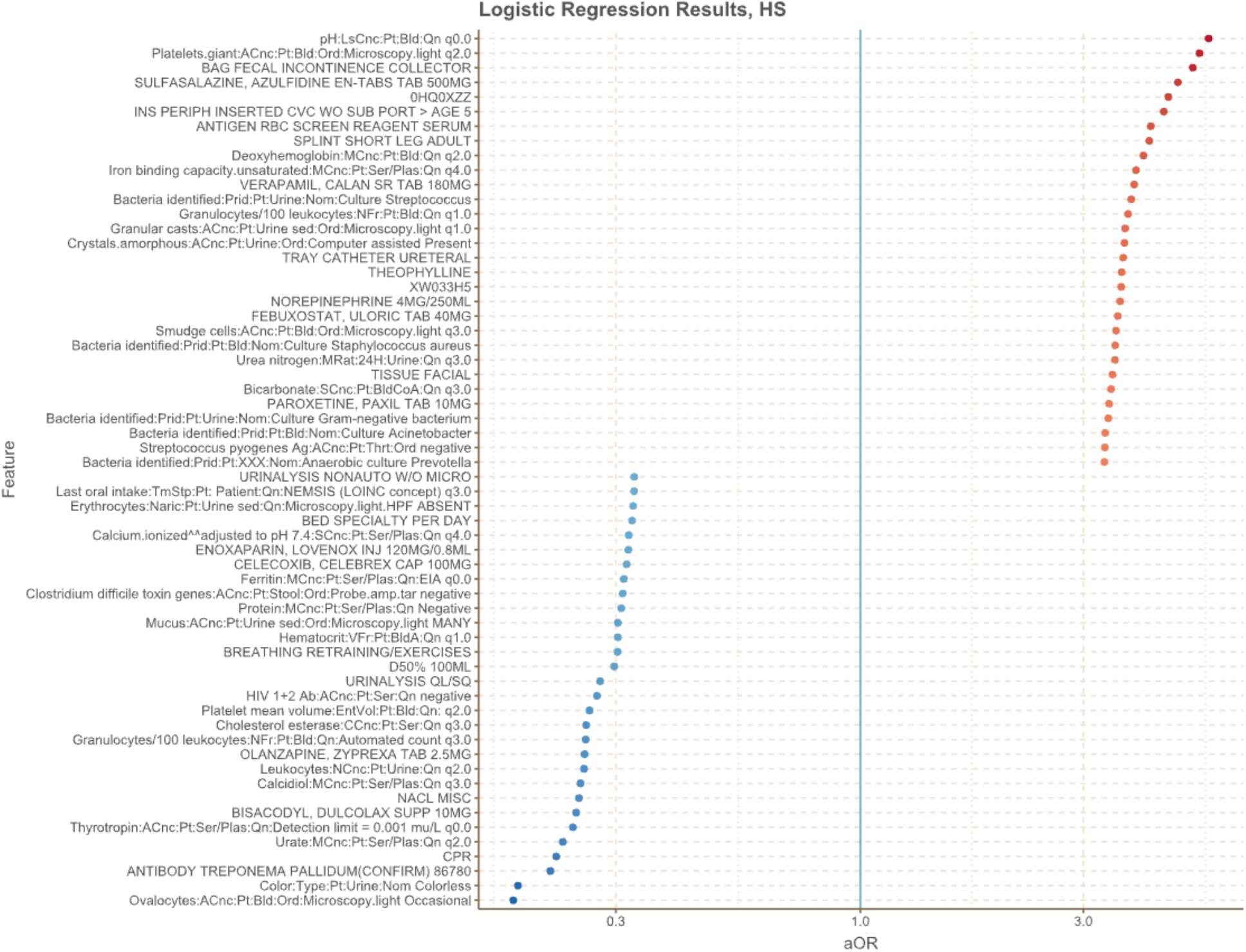
Top 30 predictive features for death, ICU admission, and hyperinflammatory syndrome from logistic regression Abbreviations: aOR, adjusted odds ratio

